# PreScription DigitaL ThErapEutic for Patients with Insomnia (SLEEP-I): A Protocol for a Pragmatic Randomized Controlled Trial

**DOI:** 10.1101/2022.02.26.22271430

**Authors:** Rachel P. Dreyer, Alyssa Berkowitz, Henry Klar Yaggi, Lynelle Schneeberg, Nilay D. Shah, Lindsay Emanuel, Bhanuprakash Kolla, Molly Jeffery, Mark Deeg, Keondae R. Ervin, Frances Thorndike, Joseph S. Ross

## Abstract

**Introduction:** Cognitive behavioral therapy for insomnia (CBT-I) is effective at treating chronic insomnia, yet in-person CBT-I can often be challenging to access. Prior studies have used technology to bridge barriers but have been unable to extensively assess the impact of the digital therapeutic on real-world patient experience and multi-dimensional outcomes. Among patients with insomnia, our aim is to determine the impact of a Prescription Digital Therapeutic (PDT) (PEAR-003b, FDA-authorized as Somryst; herein called PDT) that provides mobile-delivered CBT-I on patient-reported outcomes (PROs) and healthcare utilization.

**Methods and Analysis:** We are conducting a pragmatically designed, prospective, multi-center randomized controlled trial that leverages Hugo, a unique patient-centered health data-aggregating platform for data collection and patient follow-up from Hugo Health. A total of 100 participants with insomnia from two health centers will be enrolled onto the Hugo Health platform, provided with a linked Fitbit (Inspire 2) to track activity, and then randomized 1:1 to receive (or not) the PDT for mobile-delivered CBT-I (Somryst). The primary outcome is a change in the insomnia severity index score (ISI) score from baseline to 9-weeks post-randomization. Secondary outcomes include healthcare utilization, health utility scores, and clinical outcomes; change in sleep outcomes as measured with sleep diaries; and a change in individual PROs including depressive symptoms, daytime sleepiness, health status, stress, and anxiety. For those allocated to the PDT, we will also assess engagement with the PDT.

**Ethics and Dissemination:** The Institutional Review Boards at Yale University and the Mayo Clinic have approved the trial protocol. This trial will provide important data to patients, clinicians, and policymakers about the impact of the PDT device delivering CBT-I on PROs, clinical outcomes, and healthcare utilization. Findings will be disseminated to participants, presented at professional meetings, and published in peer-reviewed journals.

**Trial Registration Number:** NCT04909229

**Strengths and limitations of this study:**

- This is the first controlled study to examine the impact of a mobile-delivered, prescription digital therapeutic (PDT) delivering Cognitive Behavioral Therapy for Chronic Insomnia (i.e., PEAR-003b, FDA-authorized as Somryst) on real-world patients outcomes of care that includes a multi-dimensional analysis of patient benefit across guideline-recommended health domains (e.g., insomnia severity index) and healthcare utilization (e.g., emergency department visits).
- This randomized clinical trial will use Hugo, a novel patient-centered health data-aggregating platform for data collection and patient follow-up, which gathers and collates patient-reported outcomes, clinical outcomes, and healthcare utilization metrics for real-world patients with chronic insomnia. The participant has ownership over their data and contributes it to research.
- Future studies should focus on patients with chronic insomnia as well as co-morbid conditions such as major depression and whether sleep improvements can be sustained, particularly in the long-term.

## INTRODUCTION

Insomnia is one of the most prevalent health concerns and imposes a significant physical, psychological, and financial burden on patients’ lives.^1^ Up to 50% of the general adult population experience insomnia symptoms, with 12-20% meeting criteria for chronic insomnia.^2 3^ The impact on both the individual and the healthcare system is substantial. Insomnia accounts for over $100 billion of US annual healthcare costs,^4–6^ and lost productivity related to insomnia costs the US economy approximately $63 billion a year.^7^ Adults suffering from insomnia also have a higher likelihood of comorbid conditions such as depression, resulting in a reduced quality of life and higher rates of morbidity and mortality.^8^ The documented high rates and detrimental effects of insomnia and co-occurring disorders, including depression, provide a compelling rationale for identifying effective, accessible, easy-to-use, and cost-effective treatments.

There is empirical evidence indicating that cognitive-behavioral therapy for insomnia (CBT-I) can effectively treat chronic insomnia,^9–15^ including when present with co-occurring disorders like major depression,^16^ with long-lasting benefits. CBT-I is now recommended as first-line therapy for insomnia^15,17^ and its primary components include a focus on sleep restriction and consolidation, stimulus control, sleep hygiene, and cognitive restructuring.^18^ However, due to challenges associated with in-person CBT-I, such as lack of trained clinicians, poor access, and limited fidelity,^19^ attention has turned towards use of technology to overcome obstacles and deliver CBT-I interventions (e.g., Sleep Healthy Using the Internet: SHUTi).^20–22,23^ Despite promising clinical efficacy in randomized controlled trials (RCTs),^23^ these studies have been unable to rigorously assess impact of the digital therapeutic on patient experience in the real-world.

In light of this gap in knowledge, we designed the Pre**S**cription Digita**L** Th**E**rap**E**utic For **P**atients with **I**nsomnia **(SLEEP-I)** trial, a pragmatic, multi-center RCT to collect and evaluate real-world data from a mobile CBT-I prescription digital therapeutic (PEAR-003b, FDA-authorized as Somryst, herein called PDT) for patients with insomnia using Hugo,^24^ a patient-centered data aggregating platform. This approach will allow the concurrent analysis of clinical outcomes data, healthcare utilization data, and data from connected devices. The data generated will be used alongside clinically validated measures of insomnia to yield a multidimensional analysis of patient benefit. The PDT will be delivered via mobile devices to patients with insomnia as 6 treatment core CBT-I modules over 9-weeks that target three common mechanisms of action: stimulus control, sleep restriction and cognitive restructuring.^21 22 25^ We will also enroll patients in the Hugo data-aggregating platform to understand patient experience with insomnia by aggregating patients’ electronic health record (EHR) data, survey data on patient-reported outcomes (PROs), healthcare utilization metrics, and patient activity recorded via Fitbit.

## METHODS and ANALYSIS

We used the SPRINT reporting guidelines to draft this protocol paper outlining the SLEEP-I clinical trial.^26^

### Overall Study Design and Data Collection

SLEEP-I is a pragmatically-designed, prospective, multi-center RCT using a two group parallel design (PDT vs. control) by 5 assessments (baseline, 9 week, 21 week, 35 week and 61 week) to evaluate the impact of the use of the PDT on PROs and clinically validated metrics for insomnia **(Figure 1)**. This study leverages a patient-centered health data-aggregating platform called Hugo,^24^ which was initially developed to overcome many of the limitations of traditional clinical trials, such as cost and patient access.^27^ Using patients’ mobile devices or computers, Hugo aggregates electronic health data for each patient from multiple sources, including electronic health records from hospitals and physicians offices, pharmacies and payors along with data from personal digital devices and wearables, by leveraging Blue Button technology and Application Programming Interfaces. Hugo aggregates electronic health data for patients from multiple sources including EHRs (hospitals, physicians offices, clinics), pharmacies, payors, and wearables utilizing hl7 FHIR (fast healthcare interoperability resources) and other APIs (application programming interface). Hugo also has the capability of delivering patient surveys through emails or text messages which essentially enables the assessment of PROs and other information without face-to-face interaction with study coordinators after enrollment.^24^ After the consent process, all participants will be enrolled in the Hugo platform, whereby they will receive near-real time access to their electronic health data from multiple sources, which will be shared with the research team; no data will be directly obtained from healthcare systems.^28^ This study will also employ use of the Fitbit (Inspire 2) fitness tracker that integrates with Hugo to obtain multiple physiologic and sleep measurements including the number of steps per day, sleep (total sleep time in minutes), and self-reported metrics such as weight, height, and body mass index (BMI).

**Figure 1.**
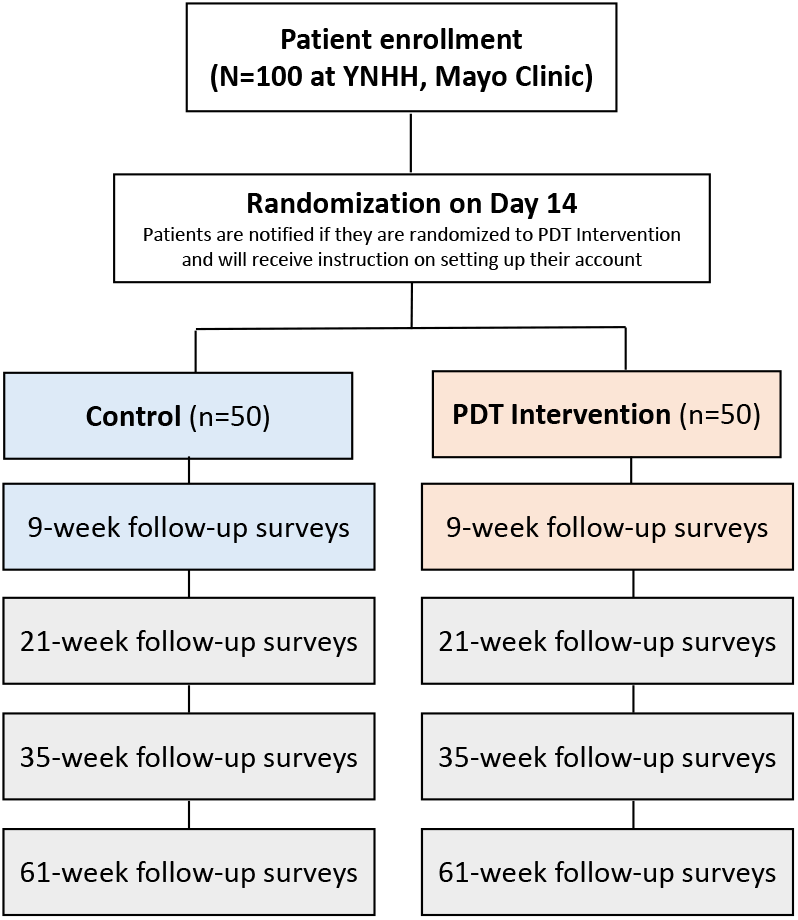
Study Design Flow of the SLEEP-I study.

### Sample Selection and Inclusion/Exclusion Criteria

Participants will be recruited from two academic health systems: Yale-New Haven Health (YNHH) and the Mayo Clinic. Potential participants with insomnia will be seen initially by a sleep provider at the YNHH and Mayo Sleep Centers. Patients with confirmed insomnia will be provided information regarding the study, and patients interested in participation will be contacted by study coordinator. Eligible patients will be consented by the study coordinator, and upon enrollment, patients will be asked to sign an electronic consent at YNHH **(Supplemental File 1)** and the Mayo Clinic **(Supplemental File 2)** as well as linking their electronic health records and Fitbit to Hugo. All participants will receive paper materials on sleep hygiene and healthy sleep tips, which include behavioral information regarding getting a good night’s sleep (e.g., setting a regular bedtime, getting out of bed if remaining awake, exercising regularly, not smoking). Participants randomized to PDT will receive access to the therapeutic for 9 weeks in addition to the sleep hygiene material.

Inclusion criteria will be confirmed prior to the informed consent process. Patients who do not meet all inclusion criteria or meet any of the non-inclusion criteria will not proceed with consent and enrollment. Patients must fulfill the following criteria prior to trial enrollment: (1) aged between 22-64 years; (2) English-speaking (both reading and writing in English required); and (3) have a diagnosis of chronic insomnia. Additionally, participants must also be willing and able to give consent to participate in the study, to have an email account (or be willing to create one), to have a smartphone capable of downloading the necessary applications and willing and able to use the PDT, the Hugo data aggregating platform and the syncable device (e.g. Fitbit).

Exclusion Criteria will include: (1) pregnancy; (2) shift work or family/other commitments that interfere with the establishment of regular night-time sleep patterns; (3) if wake/sleep time is outside the ranges of 4:00h – 10:00h (wake time) and 20:00h – 02:00h (bed-time), respectively; (4) absence of reliable internet access and smartphone; (5) a reported diagnosis of psychosis, schizophrenia or bipolar disorder or any medical disorders contraindicated with sleep restriction; (6) current involvement in a non-medication treatment program for insomnia (participants are still eligible if they are taking traditional sleep medications); (7) those with untreated co-existing sleep conditions (e.g. sleep apnea) and those who have failed CBT-I in the past.

### Intervention and Method of Assignment/Randomization

After signing informed consent documentation, participants will complete their baseline questionnaires and, over the following two weeks, will complete their baseline sleep diaries. Patients will be randomized 1:1 to the PDT or the control arm using a randomization algorithm via Hugo. A total of 100 participants with chronic insomnia from two health centers (50 at each site) will be enrolled in Hugo, provided with a linked Fitbit (Inspire 2) to track activity, and then randomized 1:1 to receive (or not) the PDT for mobile-delivered CBT-I. Patients will be notified if they are randomized to the treatment arm on day 14 by the study coordinators and will be provided with instructions on how to set up and create their PDT account if randomized to the PDT arm. The study will not employ blinding as patients will need to know if they are completing the PDT-delivered treatment.

The treatment duration will be 9 weeks, and there will be a 21-, 35-, and 61-week follow-up. All patients will be evaluated at baseline, as well as prompted to complete additional assessments at weeks 9, 21, 35, and 61 weeks post-randomization **(Figure 1)**. The PDT intervention will deliver CBT-I via mobile devices as 6 treatment core modules over 9 weeks. Using the Hugo platform, we will also collect patient-generated engagement data, healthcare utilization, and patient activity/clinical outcomes for patients with insomnia.

Patients will use their own mobile devices but will be given the necessary syncable devices to keep (i.e. Fitbit Inspire 2) and will receive a stipend for their time contributed as part of this study. This stipend will cover the consent process, initial set up and baseline questionnaire (3 hours), questionnaires provided at 9, 21, 35 and 61 weeks post-randomization, and the time it takes to sync and use the provided devices (3 hours per timepoint).

### Socio-demographic and clinical characteristics

Baseline characteristics including socio-demographic, socio-economic status, risk factors, comorbidities and sleep characteristics will be collected via self-report through Hugo at enrollment **(Table 1)**. Patients will also self-report the use of over-the-counter medications, including medications to assist with sleep and/or insomnia.

**Table 1.** Socio-demographic and clinical variables obtained at baseline from self-report.

| **Socio-demographics /**<br>**Socio-economic status** | **Risk Factors** | **Co-morbidities** | **Sleep History** |
| --- | --- | --- | --- |
| Age | Hypertension | Alcohol use | Sleep difficulties |
| Sex | Diabetes | Coronary heart disease | Insomnia treatments |
| Race/ethnicity | High cholesterol | A heart attack (also called myocardial infarction) | Length of sleep problems |
| Marital status | Smoking history | Cancer |  |
| Employment/working status |  | Depression |  |
| Education status |  | Post-Traumatic Stress Disorder (PTSD) |  |
| Annual income level |  | General Anxiety Disorder |  |
|  |  | Stroke/transient ischemic attack (TIA) |  |
|  |  | Chronic pain |  |
|  |  | Asthma or lung problems |  |

### Outcome Measures

The primary, secondary and exploratory study outcomes, including the timing of data collection/administration of measures collected through Hugo and Fitbit, are presented in **Table 2**. PROs collected in this study include the Insomnia severity index (ISI) score,^29^ the Epworth Sleepiness Scale (ESS),^30^ the Patient Health Questionnaire-8 (PHQ-8),^31^ the Generalized anxiety disorder-7 (GAD-7),^32^ the Perceived Stress Scale (PSS-10),^33^ and the Short-Form-12 (SF-12).^34^

**Table 2.** Primary, Secondary and Exploratory Outcomes.

|  |  | Weeks |  |  |  |
| --- | --- | --- | --- | --- | --- |
|  | Enrollment<br>(Baseline) | 9W | 21W | 35W | 61W |
| **Primary Outcome** |  |  |  |  |  |
| Insomnia severity index (ISI) score | x | x | x | x | x |
| **Secondary Outcomes** |  |  |  |  |  |
| **Patient-Reported Outcomes (Hugo)** |  |  |  |  |  |
| Epworth Sleepiness Scale (ESS) | x | x | x | x | x |
| Patient Health Questionnaire-8 (PHQ-8) | x | x | x | x | x |
| Generalized anxiety disorder-7 (GAD-7) | x | x | x | x | x |
| Perceived Stress Scale (PSS-10) | x | x | x | x | x |
| Short-Form-12 (SF-12) | x | x | x | x | x |
| **Healthcare Utilization Outcomes (Hugo)** |  |  |  |  |  |
| No. outpatient visits |  | x | x | x | x |
| No specialty care visits |  | x | x | x | x |
| No. medication refills for sleep |  | x | x | x | x |
| No. of medication refills for psychotropic medications |  | x | x | x | x |
| **Health Utility Scores (Hugo)** |  |  |  |  |  |
| Health utility scores (SF-12) |  | x | x | x | x |
| **Sleep Diaries\*** |  |  |  |  |  |
| Sleep efficiency (SE) | x | x | x | x | x |
| Sleep onset latency (SOL) (minutes) | x | x | x | x | x |
| Waking after sleep onset (WASO) (minutes), | x | x | x | x | x |
| Number of awakenings | x | x | x | x | x |
| Sleep quality (scale score) | x | x | x | x | x |
| Time in bed | x | x | x | x | x |
| Total sleep time | x | x | x | x | x |
| **Exploratory Outcomes (FitBit Feature Comparisons)** |  |  |  |  |  |
| Steps per day | x | x | x | x | x |
| Sleep (total sleep time in minutes) | x | x | x | x | x |
| Weight | x | x | x | x | x |
| Height | x | x | x | x | x |
| Body mass index | x | x | x | x | x |

\* Definition of key sleep outcomes:

* Definition of key sleep outcomes:

- ***Sleep efficiency* (SE):** The ratio of total sleep time (TST) to time in bed (TIB)
- **Sleep onset latency (SOL):** The length of time that it takes between “lights out” or intention to sleep and first onset of sleep
- **Sleep quality:** One’s satisfaction of the sleep experience, integrating aspects of sleep initiation, sleep maintenance, sleep quantity, and refreshment upon awakening
- **Waking after sleep onset (WASO):** Total time awake between initial sleep onset and final morning awakening

The primary outcome is a change in the ISI score^29^ from baseline to 9-weeks post-randomization. The ISI questionnaire is a 7-item global index of self-reported insomnia symptom severity that has been shown to be valid, reliable, and sensitive to changes in insomnia treatment^29 35^ and validated for online use.^36^

The secondary outcomes will be ascertained at baseline, 9-weeks, 21-weeks, 35-weeks, and 61-weeks post-randomization: (1) Healthcare utilization outcomes data available in patients’ EMR through Hugo, which may include the number of outpatient visits and specialty care visits, number of medication refills for sleep and psychotropic medications, comparing PDT to control at all follow-up time points (9-weeks, 21-weeks, 35-weeks, and 61-weeks); (2) Change from baseline to 21-, 35-, and 61-weeks in the ISI, comparing PDT to control; and (3) Change from baseline to 9-, 21-, 35-, and 61-weeks post-randomization in individual PROs including daytime sleepiness (ESS),^30^ depressive symptoms (PHQ-8),^31^ anxiety (GAD-7),^32^ stress (PSS-10)^33^ and health status (SF-12),^34^ comparing PDT to control. The ESS is the most widely used tool for measuring daytime sleepiness for clinical and research purposes.^37 38^ It is a simple, self-administered, eight-item questionnaire that measures the risk of falling asleep in eight specific situations that are commonly met. A score of less than 10 is considered normal. The higher the score (from 10 to 24), the greater the reported subjective daytime sleepiness.^30^

The PHQ-8 is a measure of depressive symptoms in the general population.^39^ Participants indicate the frequency with which they have been bothered by eight depressive symptoms (e.g., “little interest or pleasure in doing things”) in the prior two weeks. Response options range from 0 (not at all) to 3 (nearly every day) and are summed to create the total symptom severity score. The GAD-7 is a validated screening tool and measure of severity of generalized anxiety disorder,^32^ and contains seven items, with each response ranked from 0 (not at all sure) to 3 (nearly every day). A GAD-7 of 0 to 4 indicates minimal anxiety, of 5 to 9 indicates mild anxiety, 10 to 14 indicates moderate anxiety, and of 15+□indicates severe anxiety.^40^

The PSS-10 is a global perceived stress scale where respondents are evaluated on the degree to which they perceived their life situations over the past month to be unpredictable, uncontrollable, or overloaded, with higher scores indicating greater stress. Lastly, the SF-12 instrument measures overall physical and mental health status through 12 items.^41^ Both the Physical Component Summary (PCS) and Mental Component Summary (MCS) scores were used for this study and range from 0 to 100, with higher scores indicating a greater level of physical or mental functioning.

Asides from PROs administered in this study, additional secondary outcomes include (1) Change in sleep outcomes collected through sleep diaries (sleep efficiency (SE), sleep onset latency (SOL) (minutes), waking after sleep onset (WASO) (minutes), number of awakenings, sleep quality (scale score), time in bed, and total sleep time, from baseline to 9-, 21-, 35-, and 61-weeks post-randomization comparing PDT to control. Following the baseline assessment that will include questionnaires as described above, patients will complete 10 days of sleep diaries within a 14-day window as well as at all follow up time periods. The sleep diaries are part of recent guidelines from the American Academy of Sleep Medicine^17 42^ as outcomes to be considered in evaluation of efficacy and clinical significance. Other secondary outcomes will include; (2) Change in (and total) health utility scores using the SF-12 among patients randomized to receive PDT versus the control only at 9, 21, 35, and 61 weeks post-randomization. Lastly, for patients randomized to the PDT, we will examine the relationship between engagement with the PDT and clinical outcomes, particularly the sleep-specific outcomes (ISI and diary-derived sleep metrics).

The exploratory outcomes will be ascertained at 9-weeks 21-weeks, 35-weeks, and 61-weeks post-randomization. Physical and sleep activity measured using Fitbit (steps per day, sleep [total sleep time in minutes], and self-reported metrics such as weight, height, and BMI from baseline to 9-, 21-, 35-, and 61-weeks post-randomization comparing PDT to control.

### Data Analysis Plan

All analyses of results from this RCT will be conducted as intent-to-treat (ITT) to avoid the effects of crossover and dropout.^43^ We will report baseline descriptive statistics for the overall study, by site, and for both the control and treatment arms of the study. Baseline data will be compared using Pearson Chi-Squared tests or Fisher exact tests for dichotomous and/or categorical variables and student’s t-tests for continuous variables. If variables are deemed as non-parametric, we will use a median test such as a Mann–Whitney *U*-test, where appropriate.

For the primary outcome, we will use a *t*-test to compare the ISI scores^29^ for the intervention (PDT + Fitbit) and control group (Fitbit only) at baseline. We will then use a 2 (group) × 2 (time) repeated measures analysis of variance (RM ANOVAs) to compare pre to post changes from baseline to 9-weeks across groups.^21 25^ Paired sample *t* tests by group will be used to examine time effects within each condition (if the overall interaction effect is significant). At weeks 21, 35, and 61 post-randomization we will also perform the same analysis, but this will be as an exploratory secondary endpoint. If a patient drops out, we will carry forward the most recent PRO response. As this is an RCT, we expect that confounding will be minimal. If patients are missing outcome data, we will use the last observation carried forward for the patient-reported outcome. Missing covariates will be set to missing.

For the secondary outcomes using the PROs, we will calculate the change in the ESS,^30^ PHQ-8,^31^ GAD-7,^7^ PSS-10^33^ and SF-12,^34^ and the scores at baseline and at 9-weeks, 21-weeks, 35-weeks and 61-weeks post-randomization and perform a comparison between patients randomized to the intervention (PDT + Fitbit) and patients randomized to the control (Fitbit only). Based on our prior work using SHUTi,^44^ we will examine the change in scores between groups using a mixed model repeated measures ANOVA^45^ with an unstructured matrix and estimated degrees of freedom (df) with Satterthwaite’s correction. We will present df alongside F-test statistics and t statistics. For the secondary clinical/healthcare utilization outcomes we will compare the PDT to the control at all follow-up time points. These comparisons will be compared using *t*-tests at each time point.

For the sleep diary outcomes,^21^ we will calculate the change in each sleep outcome from baseline to 9-weeks, 21-weeks, 35-weeks, and 61-weeks post-randomization comparing PDT to the control based on sleep diaries. We will examine the change in scores between groups using a mixed model repeated measures ANOVA.^45^ Paired-sample *t*-tests will be used to examine time effects within each condition if the overall interaction effect is significant.

For the secondary health utilities outcome, we will calculate the change in health utility scores from baseline to 9-weeks, 21-weeks, 35-weeks, and 61-weeks post-randomization, and perform a comparison between patients randomized to the intervention (PDT + Fitbit) and patients randomized to the control (Fitbit only). Health utilities scores are derived from the SF-6D algorithm as applied to the SF-12 data.^46^

For the secondary engagement outcome, we will examine the relationship between engagement with PDT and clinical outcomes in the PDT arm at all follow-up time points. Correlations between engagement and clinical outcomes will be evaluated using both Pearson’s correlation coefficient and Spearman’s rank correlation as follows. Change from baseline (Follow up – baseline) will be calculated for ISI, sleep diary-derived metrics of SOL and WASO, and PHQ-8, at both the end of treatment and the end of all follow-ups. These will be correlated with core completion rates, sleep diary completion rate, and the number of times the PDT is opened. In addition, clinical outcomes among those who complete all six cores of treatment will be examined.

Lastly, the exploratory physical and sleep activity outcomes (measured using Fitbit) will again be compared from baseline to 9-weeks, 21-weeks, 35-weeks, and 61-weeks comparing PDT to control. We will examine the change in scores between groups using a mixed model repeated measures ANOVA.^45^ Paired-sample *t*-tests will be used to examine time effects within each condition if the overall interaction effect is significant. A value of P<0.05 will be considered statistically significant. All analyses will be done in SAS (version 9.4) and performed at the Mayo Clinic.

### Sample Size Calculation

Our sample size was determined assuming 90% power to detect an effect size of d=0.52 for the main outcome (change in ISI from baseline to 9-weeks post-randomization),^44^ with alpha 0.05 using the PASS software (PASS 15) to detect a clinically meaningful change.^47^ This effect size is 1/2 to 1/3 of what we have seen previously. Because this effect size is smaller than the levels demonstrated in randomized controlled trials, we are adequately powered to detect changes of interest in the main ISI outcome. We further note that this calculation is conservative because the analysis may optionally draw from outcome values recorded at baseline and each follow-up time. Because the models assume that each outcome is normally distributed, the outcome effects represent the average amount each outcome is expected to change with the incremental shift in any explanatory variable. We will recruit a total of 100 participants and will randomly assign them to treatment and control arms based on a 50% probability of assignment. We also assume a 25% rate of drop-out between baseline and the end of follow-up, resulting in an effective sample size of N=80.

## DISCUSSION

Knowledge gained from the SLEEP-I RCT will assist in improving the PDT which could then improve outcomes for individuals with chronic insomnia, which is one of the most common health concerns and imposes a significant burden on patients’ lives.^1^ Although CBT-I is the main treatment for insomnia, there are many challenges associated with in-person CBT-I such as poor access and lack of trained clinicians.^19^ This will be the first controlled study to address these important gaps in clinical care by examining the impact of a mobile-delivered PDT device delivering CBT for Insomnia using Hugo, a novel data science aggregating platform, to inform the field on the impact of a PDT for chronic insomnia not only on clinical domains (change in insomnia severity), but also important related domains of patient satisfaction and healthcare utilization.

Results from this study will advance our understanding of: (1) how novel ways of collecting and aggregating clinical and PROs data can support informed clinical decision-making; (2) digital therapeutic engagement and its relationship to clinical outcomes; and (3) evaluation of data from linked devices by providing novel information on a prescription digital therapeutic for insomnia, connected with the Hugo platform. The outcome of this research will provide crucial data to inform the latest thinking about how data from both digital therapeutics and EHR systems can be utilized to evaluate real-world clinical and utilization outcomes. These data will be used to demonstrate the value of implementing technology within healthcare systems, supporting the broad uptake of similar technology platforms. In addition, they will inform reimbursement discussions with payers to support coverage of and broad access to effective digital therapeutics.

This RCT has several potential study limitations. First, this study sample is quite small and will be relatively homogeneous given that participants will be recruited from sleep medicine clinics and may not represent those who only present at primary care or any medical disorders contraindicated with sleep restriction, and participants will be drawn from two urban sleep centers. Future studies should aim to enroll larger and more heterogeneous samples to improve the generalizability of the findings, such as those comparisons by sex and race/ethnicity to determine which patients most benefit from treatment, based on specific risk factors.^48–50^ Second, this study relies on participants motivation and/or willingness to complete sleep diaries/intervention cores and PRO’s. Third, our findings will be based on self-report measures or PROs (e.g., depression, stress, anxiety) versus a clinician-administered interview at all assessment points,

### Patient and public involvement

Patients and/or the public were not involved in the design, or conduct, or reporting, or dissemination plans of this research.

## Ethics and Dissemination

### Ethics Approval

The SLEEP-I RCT is sponsored by the National Evaluation System for health Technology Coordinating Center (NESTcc). Ethics approval was obtained independently at each of the two health systems, including at Yale University on August 30, 2021 (#2000029050); and Mayo Clinic on February 14, 2022 (#20-006319). Any amendments to the protocol are first reviewed by each of the two local institutional review boards prior to implementation and also receive approval from the study sponsor. This RCT is also registered at ClinicalTrials.gov (NCT04909229) and was first posted on June 1, 2021.

Serious adverse events are not expected in this study where participants will be using their own digital devices. However, if there are device-related adverse events, they will be reported immediately, followed by a written report within 5 calendar days of the PIs becoming aware of the event to the IRB (using the appropriate forms from the website) and any appropriate funding and regulatory agencies. The investigators will apprise fellow investigators and study personnel of all UPIRSOs and adverse events that occur during the conduct of this research project via email as they are reviewed by the PIs. The investigator team will make clear that any sync-able data, including PROs, will not be reviewed in real-time by researchers and will not be provided to the clinical care team and, therefore, any adverse or severe symptoms should be reported directly to their physician(s), or emergency room physicians as they would have in the normal course of care.

### Dissemination plan

The results from this RCT will be presented at both scientific meetings and submitted for publication to peer-reviewed journals. Additionally, study results will be shared with stakeholders and enrolled study participants.

## Supporting information

Supplement 1 - Yale IRB

Supplement 2 - Mayo IRB

SPIRIT Checklist

## Data Availability

All data produced in the present work are contained in the manuscript.

## Authors’ Contributions

- RPD: Conception and design, data collection, analysis and interpretation, writing the article, critical revision of the article, final approval.
- AB: Data collection, critical revision of the article, writing the article, final approval.
- HKY: Critical revision of the article, writing the article, final approval.
- LS: Critical revision of the article, writing the article, final approval.
- NS: Conception and design, analysis and interpretation, writing the article, critical revision of the article, final approval.
- LM: Data collection, critical revision of the article, writing the article, final approval.
- KBP: Critical revision of the article, writing the article, final approval.
- MJ: Critical revision of the article, writing the article, final approval.
- MD: Conception and design, data collection, analysis and interpretation, writing the article, critical revision of the article, final approval.
- KRE: Critical revision of the article, writing the article, final approval
- FT: Conception and design, data collection, analysis and interpretation, writing the article, critical revision of the article, final approval.
- JSR: Conception and design, analysis and interpretation, writing the article, critical revision of the article, final approval, overall study responsibility.

## Trial Status

This trial began on December 22, 2021.

## Data Sharing Plan

The study protocol, informed consent form, statistical analysis plan and analytic code will be made publicly available. Upon study completion, we plan to share de-identified individual patient data for use by others for research in a manner that protects patient confidentiality.

## Acknowledgments

None.

