## Supplement 1 - Yale IRB for "PreScription DigitaL ThErapEutic for Patients with Insomnia (SLEEP-I): A Protocol for a Pragmatic Randomized Controlled Trial"

### COMPOUND AUTHORIZATION AND CONSENT FOR PARTICIPATION IN A RESEARCH STUDY

#### YALE UNIVERSITY
#### YALE SCHOOL OF MEDICINE
#### YALE-NEW HAVEN HOSPITAL

**Study Title:** Randomized Controlled Trial Examining Real-World Effectiveness of a Prescription Digital Therapeutic for the Treatment of Insomnia

**Principal Investigator (the person who is responsible for this research):** Joseph Ross, MD, MHS

**Phone Number**: 203-785-2987
**24-Hour Phone Number**: 203-287-3550

**Research Study Summary:**

- We are asking you to join a research study.
- The purpose of this research study is to help us understand whether a digital Cognitive Behavioral Therapy intervention (CBT) for insomnia that is called PEAR-003b, created by Pear Therapeutics (https://peartherapeutics.com/) improves outcomes for patients with insomnia.
- Study procedures will include: Connecting your electronic health records with a patient-centered data sharing technology platform called Hugo, wearing a Fitbit, completing questionnaires at 5 different time points, and completing the digital CBT intervention (only if randomized to the intervention group).
- Only 1 visit is required.
- Your initial visit will take 2 hours maximum.
- There are some risks from participating in this study. Some questions in the questionnaires might make you feel uncomfortable. Additionally, wearing the Fitbit watch may also be uncomfortable for you, especially while you are sleeping.
- The study may have benefits to you. Knowledge gained from this study may improve outcomes for patients such as yourself who suffer from insomnia. Also, through the Hugo platform, you will have easy access to your medical records across all the institutions where you receive care. Wearing a Fitbit may also give you additional useful information regarding your health and fitness.
- Taking part in this study is your choice. You can choose to take part, or you can choose not to take part in this study. You can also change your mind at any time. Whatever choice you make, you will not lose access to your medical care or give up any legal rights or benefits.
- If you are interested in learning more about the study, please continue reading, or have someone read to you, the rest of this document. Take as much time as you need before you make your decision. Ask the study staff questions about anything you do not understand. Once you understand the study, we will ask you if you wish to participate; if so, you will have to sign this form.

**Why is this study being offered to me?**

We are asking you to take part in a research study because you are between 22-64 years of age, have a diagnosis of chronic insomnia, and have presented to the Yale-New Haven Hospital

(YNHH) Sleep Medicine Center. We are looking for 50 participants with insomnia to be part of this research study.

**Who is paying for the study?**
The Medical Device Innovation Consortium, through its National Evaluation System for health Technology Coordinating Center (NESTcc), which is funded by the U.S. Food and Drug Administration.

**What is the study about?**
The purpose of this research is for you to help us understand how we can improve outcomes for patients who have sleep problems (i.e. insomnia) by using a tailored, digital CBT intervention called PEAR-003b. Insomnia is one of the most common health concerns and imposes a significant burden on patients' lives. CBT is the main treatment for insomnia, but there are many challenges associated with in-person CBT such as lack of trained doctors, the cost and also poor access to this service. In light of this, PEAR-003b was developed. PEAR-003b is a digital CBT program that you can access on your mobile device or computer.

This study also uses a technology platform called Hugo Health that you can access on your mobile device or a computer that will gather together information (with your permission) from your online health records from your doctor's office, along with your responses to questionnaires from the researchers conducting this study and information about your activity/sleep from a Fitbit activity tracker that we will provide you.

**What are you asking me to do and how long will it take?**
If you agree to take part in this study, this is what will happen: we will obtain informed consent, collect contact information, and will randomize you into one of two groups: (1) Individuals who receive PEAR-003b (i.e. CBT-I) and Fitbit; or (2) Individuals who receive Fitbit only. By being in one of these two groups, particuarly the intervention group, this will allow us to determine upon study completion if the CBT-I improved outcomes for you or not. Both groups will be enrolled in Hugo.

Once we enroll you in the study we will ask you to complete questionnaires at 5 different time points (baseline, 9-week follow up; 21-week follow up, 35-week follow up; and 61-week follow up). These will include questions about insomnia severity, depression, stress, health-related quality of life, daytime sleepiness, and general anxiety. Both groups will receive materials on sleep hygiene and healthy sleep tips.

A description of study procedures is listed below in chronological order.

**Setup process for Hugo data sharing platform and Fitbit (Both groups)**
1. Using your own mobile device or computer, the study coordinator will help you to register for the Hugo platform. Registration for Hugo Health will require you to enter basic information including first name, last name, email address, and to choose a security password. You will then be prompted to accept standard terms and conditions and a privacy notice for the Hugo platform.
2. Using your personal mobile device (phone or tablet), you will check your email and click the confirmation link to activate your new Hugo account. If you do not have an email account and wish to create one, the study coordinator can help you set one up from a variety of free email providers.



APPROVED BY THE YALE UNIVERSITY IRB 6/3/2021

3. Once your Hugo account is confirmed, the study coordinator will then walk you through the remaining steps to complete study enrollment in the Hugo platform.
4. The study coordinator will then show you how to access and complete your enrollment questionnaire. This questionnaire will be sent to you through email or text message, depending on which you would prefer, and will link to a multiple-choice survey in a web browser for you to complete. The study coordinator will help you with any technical questions you may have when you begin the survey.
5. The study coordinator will then help you set up an account with Fitbit, including connecting the Fitbit to both your phone and the Hugo platform.
6. The Hugo platform will prompt you to link your patient portal accounts by presenting a list of participating health systems. You can select the systems where you have received care and enter your patient portal credentials (all of these are password-protected). Should you forget your password, you can request a reset link be sent to your email account. The study coordinator can assist in setting up a new YNHH MyChart account, obtain your YNHH MyChart username, and help reset your YNHH MyChart password, if needed.
7. After your health records have been linked, the Hugo platform will display your health data, which can differ for the different health systems. The study coordinator will help you with the study information and be available to answer any questions related to data sharing.
8. The study coordinator will help you with setting up accounts for other health systems, if needed.
9. You will be asked to agree to share data from Hugo with the researchers. The medical record data being shared may include medications, problems, procedures, encounters, lab results, diagnoses, vital signs, and possibly other data that become available. From the Fitbit, the data being shared may include sleep patterns, movement (steps per-day), weight and BMI.
10. At the end of this consent form, we will ask you to give the researchers permission to see health information that you connect to the Hugo platform.

***Please note:*** Researchers will not be watching or evaluating your responses to the questionnaires delivered via Hugo. None of the information collected in this study will be shared with your medical team. If at any point any medical issues arise, **please contact your doctor. In case of a life-threatening emergency, call 911 immediately.**

**Setup process for PEAR-003b App (Intervention group only)**

1. For those randomized to receive the PEAR-003b app, 14 days after enrollment the study coordinator will send you an email to your own mobile device with a registration access code as well as instructions on how to download the app from the Apple Store or Google Play.
2. You will then download and open the app using the registration access code. The app will then launch the onboarding sequence (approximately 8 introductory screens). After this is complete you can immediately begin Core 1 of the digital CBT-I treatment.
3. Core 2 will open 7 days after Core 1 is completed if you have completed at least 5 Sleep Diaries for the most recent 7 days. These will be Sleep Diaries you keep in the app. If 5 Sleep Diaries are not present, Core 2 will not open until they are complete.
4. All other Cores will open 7 days after the previous Core is completed. The study coordinator will help you with any technical questions you may have when you begin using the PEAR-003b app.



APPROVED BY THE YALE UNIVERSITY IRB 6/3/2021

**Continuous Study Process**
After the initial in-office set up is complete, you will be asked to perform the following tasks at home. If you have any questions or experience technical issues at any time, please reach out to the study coordinator's phone or email:

1. For all 63 weeks of this study, we ask that you wear your Fitbit as often as possible both during the day and while sleeping. We also ask that you turn on the Bluetooth feature on your phone at least once a week to allow your Fitbit to send data to your phone; if possible, it is preferred that you leave your Bluetooth on more often. If you are unfamiliar with the Bluetooth setting on your phone, the study coordinator will be able to help you.
2. Surveys will be sent to you throughout the duration of the follow-up period – at 9 weeks, 21 weeks, 35 weeks, and 61 weeks via a secure link sent through text or email from the Hugo platform.
3. For those in the intervention group we ask that you complete sleep diaries through the PEAR-03b app for the 9-week duration of the study. This will include information on what time you got in bed, how often you awoke during the night, and what time you got out of bed for the day.

Once the study is complete, we will remove your name and identifying information. At no point will we publish this information with individual data. Data from this study will be shared with investigators at Yale, the Mayo Clinic, and Pear Therapeutics to help us gain a better understanding about individuals with insomnia and what might lead to better outcomes. As a valued partner in this study, the information that you share will help us to learn more about the PEAR-03b CBT program and how it contributes to important patient and sleep outcomes – and whether Hugo and Fitbit can collect useful information which will inform future studies.

**What are the risks and discomforts of participating?**
While participating in this study, you will be asked to fill out multiple surveys, which will take some time and may be inconvenient. Some of the surveys may include sensitive questions that you may feel uncomfortable answering. If you feel uncomfortable answering any specific survey question, you will be able to skip these questions and continue with the rest of the survey. You may also find it inconvenient to wear the provided Fitbit device or connect it to your phone or tablet.

In following some intervention recommendations, participants may be asked to restrict sleep at certain times, which could lead them to initially feel more tired. This could potentially exacerbate the fatigue that many participants may already be experiencing.

**How will I know about new risks or important information about the study?**
If the study team learns that a participant has clinical depression during screening, or identifies a participant as having clinical depression during the study, the participant will be instructed to schedule an appoint with their primary care clinician; if the participant does not have a primary care clinician, the study team will offer to help the participant schedule an appointment with a primary care clinician at the New Haven Primary Care Consortium (NHPCC).
**How can the study possibly benefit me?**
Using the provided Fitbit, you may also gain additional awareness and information regarding your health and fitness. As a valued member of our research team, this research may benefit you directly in that knowledge gained from the results may improve outcomes for other patients

with insomnia – through the use of the PEAR-003b mobile-delivered CBT application and Hugo platform, with linkage to Fitbit.

**How can the study possibly benefit other people?**
The benefit to science and other people may include a better understanding of how we can improve outcomes of people with insomnia.

**Are there any costs to participation?**
You will not have to pay for taking part in this study. The only costs include transportation and your time coming to the enrollment study visit, along with the time required to complete electronic surveys during the study. You are also responsible for data charges that may be incurred for utilizing online features of PEAR-003b, Hugo or Fitbit when not connected to Wi-Fi.

**Will I be paid for participation?**
You will be paid for taking part in this study. You will recieve $150 compensation in total for your participation via a Visa pre-paid card provided through the email address you used to create your Hugo account. This stipend will cover the consent process, initial set up and baseline questionnaire, questionnaires provided at 9 weeks, 21 weeks, 35 weeks, and 61 weeks along with the time it takes to sync and use the provided devices. You are responsible for paying state, federal, or other taxes for the payments you receive for being in this study. Taxes are not withheld from your payments.

You will be paid $30 upon the completion of all questionnaires at each respective timepoint (i.e. Baseline, 9 weeks post-randomization, 21 weeks post-randomization, 35 weeks post-randomization, 61 weeks post-randomization), totaling $150.

**What are my choices if I decide not to take part in this study?**
Instead of participating in this study, you have some other choices.
You could:

- Get treatment without being in a study.
- Take part in another study.
- Not receive treatment for your disease.

**How will you keep my data safe and private?**
All identifiable information that is obtained in connection with this study will be treated as confidential and will be disclosed only with your permission or as permitted by U.S. or State law. Examples of information that we are legally required to disclose include abuse of a child or elderly person, or certain reportable diseases. Once you enroll in the study you are given a specific study number. Your name will not appear directly on any of the questionnaires. Records of your participation in this study will be kept protected and treated as confidential. All Yale based study computers will be password-protected. All collected study data will be de-identified within 12 months of study completion. The data will be kept in this anonymous form indefinitely. When the results of the research are published or discussed in conferences, no information will be included that would reveal your identity unless your specific consent for this activity is obtained. Representatives from the Yale Human Investigation Committee (the Committee that reviews, approves and monitors human subject research) may inspect study records during internal auditing procedures. However, they are required to keep all information confidential. When we publish the results of the research or talk about it in conferences, we will not use your name. If we want to use your name, we would ask you for your permission. We will also share



APPROVED BY THE YALE UNIVERSITY IRB 6/3/2021

information about you with other researchers for future research but we will not use your name or other identifiers. We will not ask you for any additional permission.

**<u>What Information Will You Collect About Me in this Study?</u>**
The information we are asking to use and share is called “Protected Health Information.” It is protected by a federal law called the Privacy Rule of the Health Insurance Portability and Accountability Act (HIPAA). In general, we cannot use or share your health information for research without your permission. If you want, we can give you more information about the Privacy Rule. Also, if you have any questions about the Privacy Rule and your rights, you can speak to a Yale Privacy Officer at 203-432-5919.

The specific information about you and your health that we will collect, use, and share includes:

- Demographics/socio-economic status (e.g., age, sex, ethnicity/race)
- Medical history
- Dates of admission to hospital and information about future hospital visits, procedures and medical care
- Emergency department encounters
- Medication data
- Outpatient visits
- Fitbit data (heart rate, tracking steps per day and/or exercise, sleep (total sleep time in minutes), and self-reported metrics such as weight, height and BMI)

**HIPAA**
By signing this form, you authorize the use and/or disclosure of the information described above for this research study. The purpose for the uses and disclosures you are authorizing is to ensure that the information relating to this research is available to all parties who may need it for research purposes.

All health care providers subject to HIPAA (Health Insurance Portability and Accountability Act) are required to protect the privacy of your information. The research staff at the Yale School of Medicine and Yale New Haven Hospital are required to comply with HIPAA and to ensure the confidentiality of your information. Some of the individuals or agencies listed below may not be subject to HIPAA and therefore may not be required to provide the same type of confidentiality protection. They could use or disclose your information in ways not mentioned in this form. However to better protect your health information, agreements are in place with these individuals and/or companies that require that they keep your information confidential.
This authorization to use and disclose your health information collected during your participation in this study will never expire. However, you have the right to change your preference at any point in the future. Identifiers will be removed from the identifiable private information and after such removal, the information you contribute to this study could be used for future research or to help inform regulatory actions and can be distributed to another investigator for future research studies without additional informed consent from you or a legally authorized representative. Outside investigators will not know who you are. Private information such as your name, birth date or medical record number will not be shared with them.

☐ **By checking this box, I acknowledge my contribution to science and that my de-identified data collected as part of this study may be used in future research or for regulatory purposes without further consent from me.**

**How will you use and share my information?**
We will use your information to conduct the study described in this consent form. We may share your information with:

- Representatives from Yale University, the Yale Human Research Protection Program and the Institutional Review Board (the committee that reviews, approves, and monitors research on human participants), who are responsible for ensuring research compliance. These individuals are required to keep all information confidential.
- The Principal Investigators, research staff, and collaborators at both Yale University and the Mayo Clinic, who are assisting with this study
- Hugo Health, the company that owns the data sharing platform, in accordance with its Privacy Policy
- Pear Therapeutics (https://peartherapeutics.com/), the company that owns the PEAR-003b CBT application, in accordance with its Privacy Policy
- The US Food and Drug Administration (FDA), as regulators of medical devices
- Your sleep doctor, primary care physician and/or their staff

We will do our best to make sure your information stays private. But, if we share information with people who do not have to follow the Privacy Rule, your information will no longer be protected by the Privacy Rule. Let us know if you have questions about this. However, to better protect your health information, agreements are in place with these individuals and/or companies that require that they keep your information confidential. Your information will not be used for commercial purposes.

**Why must I sign this document?**
By signing this form, you will allow researchers to use and disclose your information described above for this research study. This is to ensure that the information related to this research is available to all parties who may need it for research purposes. You always have the right to review and copy your health information in your medical record.

**What if I change my mind?**
The authorization to use and disclose your health information collected during your participation in this study will never expire. However, you may withdraw or take away your permission at any time. You may withdraw your permission by contacting the study staff by telephone or e-mail.

If you withdraw your permission, you will not be able to stay in this study but the care you get from your doctor outside this study will not change. No new health information identifying you will be gathered for this study after the date you withdraw. Information that has already been collected may still be used and given to others until the end of the research study to ensure the integrity of the study and/or study oversight. Once you withdraw, you will not receive further communication about this study. If you delete your Hugo account before your participation in this study ends, you will be automatically removed from the study.

**What if I want to refuse or end participation before the study is over?**
Taking part in this study is your choice. You can choose to take part, or you can choose not to take part in this study. You also can change your mind at any time. Whatever choice you make, you will not lose access to your medical care or give up any legal rights or benefits.



APPROVED BY THE YALE UNIVERSITY IRB 6/3/2021

We would still treat you with standard therapy or, at your request, refer you to a clinic or doctor who can offer this treatment. Not participating or withdrawing later will not harm your relationship with your own doctors or with this institution.

To withdraw from the study, you can call or e-mail a member of the research team at any time and tell them that you no longer want to take part.

**What will happen with my data if I stop participating?**
All data up to the date of withdrawl will be included in the study.

**Who should I contact if I have questions?**
Please feel free to ask about anything you don't understand. If you have questions later or if you have a research-related problem, you can email the PI (Joseph Ross at) or Research Associate (Alyssa Berkowitz at)

If you have questions about your rights as a research participant, or you have complaints about this research, you can call the Yale Institutional Review Boards at (203) 785-4688 or.

A description of this clinical trial will be available on *http://www.ClinicalTrials.gov*, as required by U.S. Law. This Web site will not include information that can identify you. At most, the Web site will include a summary of the results. You can search this Web site at any time.

**Authorization and Permission**

Your signature below indicates that you have read this consent document and that you agree to be in this study.

We will give you a copy of this form.

| | | |
|---|---|---|
| Participant Printed Name | Participant Signature | Date |
| Person Obtaining Consent Printed Name | Person Obtaining Consent Signature | Date |



APPROVED BY THE YALE UNIVERSITY IRB 6/3/2021
