## Supplement 2 - Mayo IRB for "PreScription DigitaL ThErapEutic for Patients with Insomnia (SLEEP-I): A Protocol for a Pragmatic Randomized Controlled Trial"

MAYO CLINIC

| |
|---|
| Name and Clinic Number |

**Approval Date:** **February 10, 2022**
**Not to be used after:** **February 9, 2023**

### RESEARCH PARTICIPANT CONSENT AND PRIVACY AUTHORIZATION FORM

**Study Title:** Randomized Controlled Trial Examining Real-World Effectiveness of a Prescription Digital Therapeutic for the Treatment of Insomnia and Depression

**IRB#:** 20-006319

**Principal Investigator:** Bhanu Kolla, M.D., and Colleagues

#### Key Study Information

| | |
|---|---|
| This section provides a brief summary of the study. It is important for you to understand why the research is being done and what it will involve before you decide. **Please take the time to read the entire consent form carefully and talk to a member of the research team before making your decision.** You should not sign this form if you have any questions that have not been answered. | |
| **It's Your Choice** | This is a research study. Being in this research study is your choice; you do not have to participate. If you decide to join, you can still stop at any time. You should only participate if you want to do so. You will not lose any services, benefits or rights you would normally have if you choose not to take part. |
| **Research Purpose** | The purpose of this study is to help us better understand whether a digital Cognitive Behavioral Therapy (CBT) intervention for insomnia called Somryst (herein called PEAR-003b), created by Pear Therapeutics (https://peartherapeutics.com/), improves outcomes for patients with insomnia and depression.<br><br>You have been invited to join this study because you are an adult with a diagnosis of chronic insomnia and depression, and you are visiting the Mayo Center for Sleep Medicine. |
| **What's Involved** | If you agree to participate, you will be asked to connect your electronic medical records from your Mayo Clinic Patient Online Services account, electronic medical records from any other health systems where you receive care, and information from a wearable device that we will provide to you, to a patient-centered health data |

| | |
|---|---|
| | sharing platform called Hugo. Through Hugo, you will be asked to answer short questionnaires that will be sent to you via your choice of e-mail or text at five different time points. You will also be asked to complete the digital Cognitive Behavioral Therapy (CBT) intervention (only if randomized to the intervention group PEAR-003b). You do not need to have any technical skills to participate in this study; a member of the study team will help you set up your phone and connect your accounts. |
| **Key Information** | This study uses a technology platform called Hugo that you can access on your mobile phone or other device that is connected to the internet. Hugo collects data from multiple sources to create your own personal health record (PHR) and empowers you to share that data with researchers. You will have control over which data sources you connect to Hugo and will have the option to turn off data sharing at any time. Hugo has a one-way link to your health care clinicians: it can access information but cannot add any information to your health records.<br><br>The Hugo platform, like many other personal health records, is not a covered entity; therefore, the HIPAA privacy rule does not apply to this platform. You will be required to sign a separate informed consent through Hugo. The Hugo platform does take all necessary precautions, including industry-standard encryption, to minimize privacy and security risks to personally identifiable information stored on behalf of study participants. Hugo makes publicly available its Security Statement (https://hugo.health/security), Privacy Notice (https://hugo.health/privacy-notice), and Terms of Service (https://hugo.health/terms-of-service/).<br><br>While participating in this study, you will be asked to fill out multiple questionnaires, which will take some time and may be inconvenient. You may skip any questions that you feel uncomfortable answering. You may also find it inconvenient to wear the provided wearable device or connect it to your phone.<br><br>We will provide you with a Fitbit for this study. The Fitbit will monitor your activity levels and sleep, and it will be yours to keep after the study is over.<br><br>If you choose to participate, you can change your mind at any time and withdraw from the study. |

Name and Clinic Number

**Approval Date:** February 10, 2022
**Not to be used after:** February 9, 2023

| | |
|---|---|
| **Learn More** | If you are interested in learning more about this study, read the rest of this form carefully. The information in this form will help you decide if you want to participate in this research or not. A member of our research team will talk with you about taking part in this study before you sign this form. If you have questions at any time, please ask us. |

#### Making Your Decision

Taking part in research is your decision. Take your time to decide. Feel free to discuss the study with your family, friends, and healthcare provider before you make your decision. Taking part in this study is completely voluntary and you do not have to participate.

If you decide to take part in this research study, you will sign this consent form to show that you want to take part. We will give you either a printed or electronic copy of this form to keep.

For purposes of this form, Mayo Clinic refers to Mayo Clinic in Arizona, Florida and Rochester, Minnesota; Mayo Clinic Health System; and all owned and affiliated clinics, hospitals, and entities.

MAYO CLINIC

Name and Clinic Number

**Approval Date:** **February 10, 2022**
**Not to be used after:** **February 9, 2023**

#### Contact Information

| If you have questions about … | You can contact … |
|---|---|
| ▪ Study tests and procedures<br>▪ Materials you receive<br>▪ Research-related appointments<br>▪ Research-related concern or complaint<br>▪ Research-related injuries or emergencies<br>▪ Withdrawing from the research study | **Principal Investigator(s):** Bhanu Kolla, M.D.<br>**Phone:** 507-255-9230<br><br>**Study Team Contact:** Lindsay Emanuel<br>**Phone:** (507) 422-6300<br><br>**Institution Name and Address:**<br>Mayo Clinic<br>200 First Street SW<br>Rochester, MN 55905 |
| ▪ Rights of a research participant | **Mayo Clinic Institutional Review Board (IRB)**<br>**Phone:** (507) 266-4000<br><br>**Toll-Free:** (866) 273-4681 |
| ▪ Rights of a research participant<br>▪ Any research-related concern or complaint<br>▪ Use of your Protected Health Information<br>▪ Stopping your authorization to use your Protected Health Information<br>▪ Withdrawing from the research study | **Research Subject Advocate (RSA)**<br>**(The RSA is independent of the Study Team)**<br>**Phone:** (507) 266-9372<br>**Toll-Free:** (866) 273-4681<br><br>**E-mail:** |
| ▪ Billing or insurance related to this research study | **Patient Account Services**<br><br>**Toll-Free:** (844) 217-9591 |

**Other Information:**

A description of this clinical trial will be available on http://www.ClinicalTrials.gov, as required by U.S. Law. This Web site will not include information that can identify you. At most, the Web site will include a summary of the results. You can search this Web site at any time.

#### Why are you being asked to take part in this research study?

You have been invited to join this study because you are an adult with a diagnosis of chronic insomnia and depression, and you are visiting the Mayo Center for Sleep Medicine.

#### Why is this research study being done?

The purpose of this study is to help us better understand how we can improve outcomes for patients who have sleeping problems (i.e. insomnia) and daytime impairments (e.g. depression symptoms) by using a tailored, digital Cognitive Behavioral Therapy (CBT) intervention called PEAR-003b. Insomnia is one of the most common health concerns and imposes a significant burden on patients' lives. Adults suffering from insomnia also have a higher likelihood of depression, resulting in a reduced quality-of-life and higher rates of death and disability. Cognitive Behavioral Therapy (CBT) is the main treatment for insomnia, but there are many challenges associated with in-person CBT such as lack of trained doctors, the cost, and also poor access to this service. In light of this, PEAR-003b was developed. PEAR-003b is a digital Cognitive Behavioral Therapy (CBT) program that you can access on your mobile device or computer.

This study also uses a technology platform called Hugo Health that you can access on your mobile device or computer that will gather together information (with your permission) from your online health records from your doctor's office, your responses to questionnaires from the researchers conducting this study, and information about your activity/sleep form a Fitbit activity tracker that we will provide you.

Please note: The Hugo Health and PEAR applications used in this study are not affiliated with or monitored by Mayo Clinic.

#### Information you should know

##### Who is Funding the Study?

The Food and Drug Administration is funding the study. The Food and Drug Administration will pay the institution to cover costs related to running the study.

MAYO CLINIC

**Approval Date: February 10, 2022**
**Not to be used after: February 9, 2023**

**Information Regarding Conflict of Interest:**

Your healthcare provider may be referring you to this research study. If your healthcare provider is also an investigator on this study, there is the chance that his or her responsibilities for the study could influence his or her recommendation for your participation. If you prefer, your healthcare provider will be happy to refer you to another investigator on the research study team for you to decide if you want to participate in the study and to see you for the research study activities while you are in the study.

#### How long will you be in this research study?

You will be in this study for about 15 months.

#### What will happen to you while you are in this research study?

If you agree to take part in this study, this is what will happen: we will obtain informed consent, collect contact information, and will randomize you into one of two groups: (1) individuals who receive PEAR-003b (i.e. CBT-I) and Fitbit; or (2) individuals who receive Fitbit only. By being in one of these two groups, particularly the intervention group, this will allow us to determine upon study completion if the CBT-I improved outcomes for you or not.

Once we enroll you in the study, we will ask you to complete questionnaires at 5 different time points (baseline, 9-week follow up; 21-week follow up, 35-week follow up; and 61 week follow up). These will include questions about insomnia severity, depression, stress, health related quality of life, daytime sleepiness, and general anxiety. Both groups will receive materials on sleep hygiene and healthy sleep tips.
A description of study procedures is listed below in chronological order.

**Setup process for Hugo data sharing platform and Fitbit (both groups)**

1. Using your own mobile device or computer, the study coordinator will help you to register for the Hugo platform. Registration for Hugo Health will require you to enter basic information including first name, last name, email address, and to choose a security password. You will then be prompted to accept standard terms and conditions and a privacy notice for the Hugo platform.
2. Using your personal mobile device (phone or tablet), you will check your email and click the confirmation link to activate your new Hugo account. If you do not have an email

account and wish to create one, the study coordinator can help you set one up from a variety of free email providers.
3. Once your Hugo account is confirmed, the study coordinator will then walk you through the remaining steps to complete study enrollment in the Hugo platform.
4. The study coordinator will then show you how to access and complete your enrollment questionnaire. This questionnaire will be sent to you through email or text message, depending on which you would prefer, and will link to a multiple-choice survey in a web browser for you to complete. The study coordinator will help you with any technical questions you may have when you begin the survey.
5. The study coordinator will then help you set up an account with Fitbit, including connecting the Fitbit to both your phone and the Hugo platform.
6. The Hugo platform will prompt you to link your patient portal accounts by presenting a list of participating health systems. You can select the systems where you have received care and enter your patient portal credentials (all of these are password-protected). Should you forget your password, you can request a reset link be sent to your email account. The study coordinator can assist in setting up a new Mayo Clinic patient portal account, obtain your Mayo Clinic patient portal username, and help reset your Mayo Clinic patient portal password, if needed.
7. After your health records have been linked, the Hugo platform will display your health data, which can differ for the different health systems. The study coordinator will help you with the study information and be available to answer any questions related to data sharing.
8. The study coordinator will help you with setting up accounts for other health systems, if needed.
9. You will be asked to agree to share data from Hugo with the researchers. The medical record data being shared may include medications, problems, procedures, encounters, lab results, diagnoses, vital signs, and possibly other data that becomes available. From the Fitbit, the data being shared may include sleep patterns, movement (steps per-day), weight and BMI.
10. At the end of this consent form, we will ask you to give the researchers permission to see health information that you connect to the Hugo platform.
11. On day 14 is when you will be randomized. You will be randomized 1:1 to the digital therapeutic or the control arm by the study coordinator using a randomization algorithm via Hugo. You will be notified if you are randomized to the treatment arm on day 14 by the study coordinator and will be provided with instruction on how to set up and create their Pear-003b account if randomized to the Pear-003b arm.

***Please note:*** Researchers will not be watching or evaluating your responses to the questionnaires delivered via Hugo. None of the information collected in this study will be shared with your medical team. If at any point any medical issues arise, **please contact your doctor. In case of a life-threatening emergency, call 911 immediately.**

**If you are unable to complete enrollment at this time, additional time can be scheduled within the next 3 days to complete the remaining steps.*

**<u>Setup process for PEAR-003b App (intervention group only)</u>**

1. For those randomized to receive the PEAR-003b app, 14 days after enrollment the study coordinator will send you an email to your own mobile device with a registration access code as well as instructions on how to download the app from the Apple Store or Google Play.
2. You will then download and open the app using the registration access code. The app will then launch in the onboarding sequence (approximately 8 introductory screens). After this is complete you can immediately begin Core 1 module of the digital CBT-I treatment.
3. Core 2 module will open 7 days after Core 1 module is completed if you have completed at least 5 Sleep Diaries for the most recent 7 days. These will be Sleep Diaries you keep in the app. If 5 Sleep Diaries are not present, Core 2 module will not open until they are complete.
4. All other Cores (modules) will open 7 days after the previous Core (module) is completed. The study coordinator will help you with any technical questions you may have when you begin using the PEAR-003b app.

   Each module (Core) includes weekly Insomnia Severity Index assessment, sleep diaries, and a learning focus. The PHQ8 is also included at the beginning of Cores 1, 3, and 5. There is a Self Assessment of patient sleep problems and Goal Setting selection in Core 1. Core 6 includes a final Self Assessment and Goals assessment to review progress made in the treatment. Each module is a combination of written material, expert explanation videos, interactions, and vignettes (written and video) of "typical" users and their journeys. Each module (Core) focuses on a different aspect of sleep behaviors and strategies

*Continuous Study Process*

After the initial in-office set up is complete, you will be asked to perform the following tasks at home. If you have any questions or experience technical issues at any time, please reach out to the PI or study coordinator's phone or email:

- For all 63 weeks of this study, we ask that you wear your Fitbit as often as possible both during the day and while sleeping. We also ask that you turn on the Bluetooth feature on your phone at least once a week to allow your Fitbit to send data to your phone; if possible, it is preferred that you leave your Bluetooth on more often. If you are unfamiliar with the Bluetooth setting on your phone, the study coordinator will be able to help you.
- Surveys will be sent to you throughout the duration of the follow-up period – at 9 weeks, 21 weeks, 35 weeks, and 61 weeks via a secure link sent through text or email from the Hugo platform.

- The study coordinator will contact you at weeks 35 and 61 to ask about care at other health systems and the Hugo platform will be checked to ensure that data from those health systems are included. You will be asked to link those health systems or provide data, as appropriate.
- For those in the intervention group (PEAR-003b) we ask that you complete sleep diaries through the PEAR-03b app for the 9-week duration of the study._This will include information on what time you got in bed, how often you awoke during the night, and what time you got out of bed for the day.

Once the study is complete, we will remove your name and identifying information. At no point will we publish this information with individual data. Data from this study will be shared with investigators at Yale, the Mayo Clinic, and Pear Therapeutics to help us gain a better understanding about individuals with insomnia and depression and what might lead to better outcomes. As a valued partner in this study, the information that you share will help us to learn more about the PEAR-03b_CBT program and how it contributes to important patient and sleep outcomes – and whether Hugo and Fitbit can collect useful information which will inform future studies.

---

#### What are the possible risks or discomforts from being in this research study?

---

As with all research, there is a chance that confidentiality could be compromised; however, we take precautions to minimize this risk.

The risk to your privacy is that the Hugo platform collects personally identifiable information (like your name and where you go to the doctor) and protected health information (like the conditions you have and medications you take). The Hugo platform is not considered a "covered entity" under the Health Insurance Portability and Accountability Act of 1996 (HIPAA); this means that the HIPAA privacy rule does not apply to this platform. The Hugo platform does take all necessary precautions, including industry-standard encryption, to minimize privacy and security risks to your stored personally identifiable information. To learn more about Hugo's commitment to the security and privacy of your data, you can visit the following links: Security Statement (https://hugo.health/security), Privacy Notice (https://hugo.health/privacy-notice), Terms of Service (https://hugo.health/terms-of-service/).

While participating in this study, you will be asked to fill out multiple surveys, which will take some time and may be inconvenient. Some of the surveys may include questions that you feel uncomfortable answering. If you feel uncomfortable answering any specific survey question, you will be able to skip these questions and continue with the rest of the survey. You may also find it inconvenient to wear the provided Fitbit device or connect it to your phone.

In following some intervention recommendations, participants may be asked to restrict sleep at certain times, which could lead them to initially feel more tired. This could potentially exacerbate the fatigue that many participants may already be experiencing.

#### Are there reasons you might leave this research study early?

You may decide to stop at any time.

In addition, the Principal Investigator or Mayo Clinic may stop you from taking part in this study at any time if it is in your best interest, if you don't follow the study procedures, or if the study is stopped.

If you leave this research study early, or are withdrawn from the study, no more information about you will be collected; however, information already collected about you in the study may continue to be used.

We will tell you about any new information that may affect your willingness to stay in the research study.

#### What if you are injured from your participation in this research study?

**Where to get help:**

If you think you have suffered a research-related injury, you should promptly notify the Principal Investigator listed in the Contact Information at the beginning of this form. Mayo Clinic will offer care for research-related injuries, including first aid, emergency treatment and follow-up care as needed.

**Who will pay for the treatment of research related injuries:**

Care for such research-related injuries will be billed in the ordinary manner, to you or your insurance. Treatment costs for research-related injuries not covered by your insurance will be paid by Mayo Clinic.

#### What are the possible benefits from being in this research study?

A possible benefit of this study is that you will have easy access to the information contained in your Mayo Clinic and outside health records that you connect to Hugo Health. Using the provided Fitbit, you may also gain additional awareness and information regarding your health and fitness. The knowledge gained from the results may improve outcomes for other patients with insomnia and depression through the use of the PEAR-003b mobile-delivered CBT application and Hugo platform with linkage to Fitbit.

#### What alternative do you have if you choose not to participate in this research study?

If you decide not to participate in this study, you will still have access to medical care and to your medical records as you would normally. You may decline to participate in the study for any reason without affecting your medical care.

#### What tests or procedures will you need to pay for if you take part in this research study?

The platform, technology, devices and applications used in this study will be provided to participants free of cost. Updates to the platform will also be provided free of cost for the duration of the study.

Participants will still be responsible for any costs associated with transportation to come to the enrollment study visit and routine follow up or health care visits that occur in the context of standard care. Participants will still be responsible for any co-payments required by their insurance company for standard treatments as well.

Participants are responsible for data charges that may be incurred from using online features of the Hugo or smart watch mobile applications when not connected to Wi-Fi.

**If you have billing or insurance questions call Patient Account Services at the telephone number provided in the Contact Information section of this form.**

#### Will you be paid for taking part in this research study?

You will be paid for taking part in this study. You will receive. $150 compensation in total for your participation via a Visa pre-paid card provided through the email address you used to create your Hugo account. This stipend will cover the consent process, initial set up and baseline questionnaire, questionnaires provided at 9 weeks, 21 weeks, 35 weeks, and 61 weeks along with the time it takes to sync and use the provided devices. You are responsible for paying state, federal, or other taxes for the payments you receive for being in this study. Taxes are not withheld from your payments.

You will be paid $30 upon the completion of all questionnaires at each respective timepoint (i.e. Baseline, 9 weeks post-randomization, 21 weeks post-randomization, 35 weeks post-randomization, 61 weeks post-randomization).

#### Will your information or samples be used for future research?

Identifiable information such as your name, Mayo Clinic number, or date of birth may be removed from your information or samples collected in this study, allowing the information or samples to be used for future research or shared with other researchers without your additional informed consent.

Identifiable data through the Hugo dashboard will be viewable by our collaborators from Yale New Haven for monitoring and compliance reasons.

#### How will your privacy and the confidentiality of your records be protected?

Mayo Clinic is committed to protecting the confidentiality of information obtained about you in connection with this research study.

During this research, information about your health will be collected. Under Federal law called the Privacy Rule, health information is private. However, there are exceptions to this rule, and you should know who may be able to see, use and share your health information for research and why they may need to do so. Information about you and your health cannot be used in this research study without your written permission. If you sign this form, it will provide that permission (or “authorization”) to Mayo Clinic.

##### Your health information may be collected from:

- Past, present and future medical records at Mayo Clinic and all health systems that you import into Hugo.
- Data collected from the provided Fitbit during the 63-week follow-up period.
- The Hugo platform (including pharmacy records and imported claims using CMS Blue Button).
- Research procedures, such as phone calls, e-mails, and research questionnaires as part of this research.

##### Your health information will be used and/or given to others to:

- Do the research.
- Report the results.
- See if the research was conducted following the approved study plan, and applicable rules and regulations.

##### Your health information may be used and shared with:

- Mayo Clinic research staff involved in this study.
- Researchers involved in this study at other institutions (Yale-New Haven Hospital, Pear Therapeutics, and Hugo Health).
- The sponsor(s) of this study and the people or groups hired by the sponsor(s) to help perform this research.
- The Mayo Clinic Institutional Review Board that oversees the research.
- Federal and State agencies (such as the Food and Drug Administration, the Department of Health and Human Services, the National Institutes of Health and other United States agencies) or government agencies in other countries that oversee or review research.
- A group that oversees the data (study information) and safety of this research.

The data collected in your Hugo account, including data from any portals you connect and responses to any questionnaires you complete, will not be transferred back to your medical record. This means that your doctors will not see your responses to the study questionnaires or the information from your wearable device.

All health care providers subject to HIPAA (Health Insurance Portability and Accountability Act) are required to protect the privacy of your information. The research staff at Mayo Clinic is required to comply with HIPAA and to ensure the confidentiality of your information. Some of the individuals or agencies listed above may not be subject to HIPAA and, therefore, may not be required to provide the same type of confidentiality protection. They could use or disclose your information in ways not mentioned in this form. However, to better protect your health information, agreements are in place with these individuals and/or companies that require that they keep your information confidential. In addition, even though Hugo Health is not required to comply with HIPAA, they maintain the highest standards of confidentiality and security of your information and will never share your data beyond this study without your expressed explicit permission as described in their privacy notice provided when you sign up for Hugo.

##### How your information may be shared with others:

While taking part in this study, you will be assigned a code that is unique to you, but does not include information that directly identifies you. This code will be used if your study information is sent outside of Mayo Clinic. The groups or individuals who receive your coded information will use it only for the purposes described in this consent form.

If the results of this study are made public (for example, through scientific meetings, reports or media), information that identifies you will not be used.

In addition, individuals involved in study oversight and not employed by Mayo Clinic may be allowed to review your health information included in past, present, and future medical and/or research records. This review may be done on-site at Mayo Clinic or remotely (from an off-site location). These records contain information that directly identifies you. However, the individuals will not be allowed to record, print, or copy (using paper, digital, photographic or other methods), or remove your identifying information from Mayo Clinic.

##### Is your health information protected after it has been shared with others?

Mayo Clinic asks anyone who receives your health information from us to protect your privacy; however, once your information is shared outside Mayo Clinic, we cannot promise that it will remain private and it may no longer be protected by the Privacy Rule.

---

#### Your Rights and Permissions

---

Participation in this study is completely voluntary. You have the right not to participate at all. Even if you decide to be part of the study now, you may change your mind and stop at any time. You do not have to sign this form, but if you do not, you cannot take part in this research study.

Deciding not to participate or choosing to leave the study will not result in any penalty. Saying ‘no’ will not harm your relationship with your own doctors or with Mayo Clinic.

If you cancel your permission for Mayo Clinic to use or share your health information, your participation in this study will end and no more information about you will be collected; however, information already collected about you in the study may continue to be used.

You can cancel your permission for Mayo Clinic to use or share your health information at any time by sending a letter to the address below:

Mayo Clinic
Office for Human Research Protection
ATTN: Notice of Revocation of Authorization
201 Building 4-60
200 1st Street SW
Rochester, MN 55905

Please be sure to include in your letter or email:
- The name of the Principal Investigator,
- The study IRB number and /or study name, and
- Your contact information.

Your permission for Mayo Clinic to use and share your health information lasts forever, unless you cancel it.

There is no expiration or end date related to the Sponsor's use of your health information received from Mayo Clinic as part of this study.

MAYO CLINIC

Name and Clinic Number

**Approval Date:** **February 10, 2022**
**Not to be used after:** **February 9, 2023**

#### Enrollment and Permission Signatures

**Your signature documents your permission to take part in this research.**

| | / / | : AM/PM |
|---|---|---|
| Printed Name | Date | Time |

Signature

**Person Obtaining Consent**

- I have explained the research study to the participant.
- I have answered all questions about this research study to the best of my ability.

| | / / | : AM/PM |
|---|---|---|
| Printed Name | Date | Time |

Signature
