## Supplementary material for "PreScription DigitaL ThErapEutic for Patients with Insomnia (SLEEP-I): A Protocol for a Pragmatic Randomized Controlled Trial": SPIRIT Checklist

Reporting checklist for protocol of a clinical trial.

Based on the SPIRIT guidelines.

**Instructions to authors**

Complete this checklist by entering the page numbers from your manuscript where readers will find each of the items listed below.

Your article may not currently address all the items on the checklist. Please modify your text to include the missing information. If you are certain that an item does not apply, please write "n/a" and provide a short explanation.

Upload your completed checklist as an extra file when you submit to a journal.

In your methods section, say that you used the SPIRIT reporting guidelines, and cite them as:

Chan A-W, Tetzlaff JM, Altman DG, Laupacis A, Gøtzsche PC, Krleža-Jerić K, Hróbjartsson A, Mann H, Dickersin K, Berlin J, Doré C, Parulekar W, Summerskill W, Groves T, Schulz K, Sox H, Rockhold FW, Rennie D, Moher D. SPIRIT 2013 Statement: Defining standard protocol items for clinical trials. Ann Intern Med. 2013;158(3):200-207

|  | | Reporting Item | Page  Number |
| --- | --- | --- | --- |
| **Administrative information** |  |  |  |
| Title | [#1](https://www.goodreports.org/spirit/info/#1) | Descriptive title identifying the study design, population, interventions, and, if applicable, trial | 1 |
|  |  | acronym |  |
| Trial registration | [#2a](https://www.goodreports.org/spirit/info/#2a) | Trial identifier and registry name. If not yet registered, name of intended registry | 4 |
| Trial registration: data set | [#2b](https://www.goodreports.org/spirit/info/#2b) | All items from the World Health Organization Trial Registration Data Set | 1-19 |
| Protocol version | [#3](https://www.goodreports.org/spirit/info/#3) | Date and version identifier | 1 |
| Funding | [#4](https://www.goodreports.org/spirit/info/#4) | Sources and types of financial, material, and other | 22 |
|  |  | support |  |

Roles and responsibilities: contributorship

[#5a](https://www.goodreports.org/spirit/info/#5a)	Names, affiliations, and roles of protocol contributors	1-2

Roles and responsibilities: sponsor contact information

[#5b](https://www.goodreports.org/spirit/info/#5b)	Name and contact information for the trial sponsor	1-2

Roles and responsibilities: sponsor and funder

[#5c](https://www.goodreports.org/spirit/info/#5c)	Role of study sponsor and funders, if any, in study	18, 22

design; collection, management, analysis, and interpretation of data; writing of the report; and the decision to submit the report for publication, including whether they will have ultimate authority over any of these activities

Roles and responsibilities: committees

[#5d](https://www.goodreports.org/spirit/info/#5d)	Composition, roles, and responsibilities of the	NA coordinating centre, steering committee, endpoint

adjudication committee, data management team, and other individuals or groups overseeing the trial, if applicable (see Item 21a for data monitoring committee)

### Introduction

Background and rationale

[#6a](https://www.goodreports.org/spirit/info/#6a)	Description of research question and justification for	6

undertaking the trial, including summary of relevant studies (published and unpublished) examining benefits and harms for each intervention

Background and rationale: choice of comparators

[#6b](https://www.goodreports.org/spirit/info/#6b)	Explanation for choice of comparators	6

Objectives	[#7](https://www.goodreports.org/spirit/info/#7)	Specific objectives or hypotheses	7

Trial design	[#8](https://www.goodreports.org/spirit/info/#8)	Description of trial design including type of trial (eg,	6

parallel group, crossover, factorial, single group), allocation ratio, and framework (eg, superiority, equivalence, non-inferiority, exploratory)

### Methods: Participants,

| **interventions, and** |  | | |
| --- | --- | --- | --- |
| **outcomes** |  |  |  |
| Study setting | [#9](https://www.goodreports.org/spirit/info/#9) | Description of study settings (eg, community clinic, | 7 |
|  |  | academic hospital) and list of countries where data |  |
|  |  | will be collected. Reference to where list of study sites can be obtained |  |
| Eligibility criteria | [#10](https://www.goodreports.org/spirit/info/#10) | Inclusion and exclusion criteria for participants. If | 8-10 |
|  |  | applicable, eligibility criteria for study centres and  individuals who will perform the interventions (eg, |  |
|  |  | surgeons, psychotherapists) |  |
| Interventions: description | [#11a](https://www.goodreports.org/spirit/info/#11a) | Interventions for each group with sufficient detail to allow replication, including how and when they will be | 10-11 |
|  |  | administered |  |
| Interventions: modifications | [#11b](https://www.goodreports.org/spirit/info/#11b) | Criteria for discontinuing or modifying allocated interventions for a given trial participant (eg, drug | NA |
|  |  | dose change in response to harms, participant |  |
|  |  | request, or improving / worsening disease) |  |
| Interventions: | [#11c](https://www.goodreports.org/spirit/info/#11c) | Strategies to improve adherence to intervention | N/A |
| adherence |  | protocols, and any procedures for monitoring adherence (eg, drug tablet return; laboratory tests) |  |
| Interventions: | [#11d](https://www.goodreports.org/spirit/info/#11d) | Relevant concomitant care and interventions that are | 8-10 |
| concomitant care |  | permitted or prohibited during the trial |  |
| Outcomes | [#12](https://www.goodreports.org/spirit/info/#12) | Primary, secondary, and other outcomes, including | 11-13 |
|  |  | the specific measurement variable (eg, systolic blood |  |
|  |  | pressure), analysis metric (eg, change from baseline,  final value, time to event), method of aggregation (eg, |  |
|  |  | median, proportion), and time point for each outcome. |  |
|  |  | Explanation of the clinical relevance of chosen efficacy and harm outcomes is strongly recommended |  |
| Participant timeline | [#13](https://www.goodreports.org/spirit/info/#13) | Time schedule of enrolment, interventions (including | 9-11 |
|  |  | any run-ins and washouts), assessments, and visits  for participants. A schematic diagram is highly |  |
|  |  | recommended (see Figure) |  |
| Sample size | [#14](https://www.goodreports.org/spirit/info/#14) | Estimated number of participants needed to achieve | 16 |
|  |  | study objectives and how it was determined, including |  |

clinical and statistical assumptions supporting any sample size calculations

Recruitment	[#15](https://www.goodreports.org/spirit/info/#15)	Strategies for achieving adequate participant

enrolment to reach target sample size

10-11

### Methods: Assignment of interventions (for controlled trials)

Allocation: sequence generation

[#16a](https://www.goodreports.org/spirit/info/#16a)	Method of generating the allocation sequence (eg,	10

computer-generated random numbers), and list of any factors for stratification. To reduce predictability of a random sequence, details of any planned restriction (eg, blocking) should be provided in a separate document that is unavailable to those who enrol participants or assign interventions

Allocation concealment mechanism

[#16b](https://www.goodreports.org/spirit/info/#16b)	Mechanism of implementing the allocation sequence	10

(eg, central telephone; sequentially numbered, opaque, sealed envelopes), describing any steps to conceal the sequence until interventions are assigned

Allocation: implementation

[#16c](https://www.goodreports.org/spirit/info/#16c)	Who will generate the allocation sequence, who will	10

enrol participants, and who will assign participants to interventions

Blinding (masking)	[#17a](https://www.goodreports.org/spirit/info/#17a) Who will be blinded after assignment to interventions	NA

(eg, trial participants, care providers, outcome assessors, data analysts), and how

Blinding (masking): emergency unblinding

[#17b](https://www.goodreports.org/spirit/info/#17b) If blinded, circumstances under which unblinding is	NA permissible, and procedure for revealing a

participant’s allocated intervention during the trial

### Methods: Data collection, management, and analysis

Data collection plan	[#18a](https://www.goodreports.org/spirit/info/#18a) Plans for assessment and collection of outcome,

baseline, and other trial data, including any related processes to promote data quality (eg, duplicate

11-14

|  | | measurements, training of assessors) and a |  |
| --- | --- | --- | --- |
|  |  | description of study instruments (eg, questionnaires,  laboratory tests) along with their reliability and validity, |  |
|  |  | if known. Reference to where data collection forms |  |
|  |  | can be found, if not in the protocol |  |
| Data collection plan: retention | [#18b](https://www.goodreports.org/spirit/info/#18b) | Plans to promote participant retention and complete follow-up, including list of any outcome data to be | 11-14 |
|  |  | collected for participants who discontinue or deviate |  |
|  |  | from intervention protocols |  |
| Data management | [#19](https://www.goodreports.org/spirit/info/#19) | Plans for data entry, coding, security, and storage, | 11-14 |
|  |  | including any related processes to promote data |  |
|  |  | quality (eg, double data entry; range checks for data  values). Reference to where details of data |  |
|  |  | management procedures can be found, if not in the |  |
|  |  | protocol |  |
| Statistics: outcomes | [#20a](https://www.goodreports.org/spirit/info/#20a) | Statistical methods for analysing primary and | 11-15 |
|  |  | secondary outcomes. Reference to where other |  |
|  |  | details of the statistical analysis plan can be found, if not in the protocol |  |
| Statistics: additional | [#20b](https://www.goodreports.org/spirit/info/#20b) | Methods for any additional analyses (eg, subgroup | 13-16 |
| analyses |  | and adjusted analyses) |  |
| Statistics: analysis | [#20c](https://www.goodreports.org/spirit/info/#20c) | Definition of analysis population relating to protocol | 11-15 |
| population and  missing data |  | non-adherence (eg, as randomised analysis), and any  statistical methods to handle missing data (eg, |  |
|  |  | multiple imputation) |  |
| **Methods: Monitoring** |  |  |  |
| Data monitoring: formal committee | [#21a](https://www.goodreports.org/spirit/info/#21a) | Composition of data monitoring committee (DMC); summary of its role and reporting structure; statement | NA |
|  |  | of whether it is independent from the sponsor and |  |
|  |  | competing interests; and reference to where further |  |
|  |  | details about its charter can be found, if not in the  protocol. Alternatively, an explanation of why a DMC |  |
|  |  | is not needed |  |

Data monitoring: interim analysis

[#21b](https://www.goodreports.org/spirit/info/#21b) Description of any interim analyses and stopping	NA guidelines, including who will have access to these

interim results and make the final decision to terminate the trial

Harms	[#22](https://www.goodreports.org/spirit/info/#22)	Plans for collecting, assessing, reporting, and managing solicited and spontaneously reported adverse events and other unintended effects of trial interventions or trial conduct

13-14

Auditing	[#23](https://www.goodreports.org/spirit/info/#23)	Frequency and procedures for auditing trial conduct, if	NA

any, and whether the process will be independent from investigators and the sponsor

### Ethics and dissemination

Research ethics approval

[#24](https://www.goodreports.org/spirit/info/#24)	Plans for seeking research ethics committee /	18

institutional review board (REC / IRB) approval

Protocol amendments	[#25](https://www.goodreports.org/spirit/info/#25)	Plans for communicating important protocol	NA

modifications (eg, changes to eligibility criteria, outcomes, analyses) to relevant parties (eg, investigators, REC / IRBs, trial participants, trial registries, journals, regulators)

Consent or assent	[#26a](https://www.goodreports.org/spirit/info/#26a)	Who will obtain informed consent or assent from	10

potential trial participants or authorised surrogates, and how (see Item 32)

Consent or assent: ancillary studies

[#26b](https://www.goodreports.org/spirit/info/#26b) Additional consent provisions for collection and use of	NA participant data and biological specimens in ancillary

studies, if applicable

Confidentiality	[#27](https://www.goodreports.org/spirit/info/#27)	How personal information about potential and enrolled

participants will be collected, shared, and maintained in order to protect confidentiality before, during, and after the trial

18-19

Declaration of interests

[#28](https://www.goodreports.org/spirit/info/#28)	Financial and other competing interests for principal	22-23

investigators for the overall trial and each study site

Data access	[#29](https://www.goodreports.org/spirit/info/#29)	Statement of who will have access to the final trial

dataset, and disclosure of contractual agreements that limit such access for investigators

13-14

Ancillary and post trial care

[#30](https://www.goodreports.org/spirit/info/#30)	Provisions, if any, for ancillary and post-trial care, and	NA for compensation to those who suffer harm from trial

participation

Dissemination policy: trial results

[#31a](https://www.goodreports.org/spirit/info/#31a)	Plans for investigators and sponsor to communicate	18-19

trial results to participants, healthcare professionals, the public, and other relevant groups (eg, via publication, reporting in results databases, or other data sharing arrangements), including any publication restrictions

Dissemination policy: authorship

[#31b](https://www.goodreports.org/spirit/info/#31b) Authorship eligibility guidelines and any intended use	NA of professional writers

Dissemination policy: reproducible research

[#31c](https://www.goodreports.org/spirit/info/#31c)	Plans, if any, for granting public access to the full	NA protocol, participant-level dataset, and statistical code

### Appendices

Informed consent materials

[#32](https://www.goodreports.org/spirit/info/#32)	Model consent form and other related documentation given to participants and authorised surrogates

Supplement 1-2

Biological specimens	[#33](https://www.goodreports.org/spirit/info/#33)	Plans for collection, laboratory evaluation, and	NA

storage of biological specimens for genetic or molecular analysis in the current trial and for future use in ancillary studies, if applicable

The SPIRIT checklist is distributed under the terms of the Creative Commons Attribution License CC- BY-ND 3.0. This checklist was completed on 05. May 2020 using <https://www.goodreports.org/>, a tool made by the [EQUATOR Network](https://www.equator-network.org/) in collaboration with [Penelope.ai](https://www.penelope.ai/)
